# Proteogenomics of glioblastoma associates molecular patterns with survival

**DOI:** 10.1101/2020.04.28.20083501

**Authors:** Gali Yanovich-Arad, Paula Ofek, Eilam Yeini, Artem Danilevsky, Noam Shomron, Rachel Grossman, Ronit Satchi-Fainaro, Tamar Geiger

## Abstract

Glioblastoma (GBM) is the most aggressive form of glioma, with poor prognosis exhibited by most patients, and a median survival time of less than two years. To examine survival-associated patterns, we assembled a cohort of 87 GBM patients whose survival ranges from less than 3 months and up to 10 years, most of which are not bearing isocitrate-dehyderogenase (IDH)-1 mutation and did not undergo prior treatment. We integrated high-resolution mass-spectrometry proteomics and RNA-sequencing to examine the yet unresolved proteomic contribution to poor patient outcome, and compared it to the more established transcriptomic contribution and to published single-cell RNA-sequencing data. Discovering both layer-specific and shared processes, we found that immune, metabolic and developmental processes distinguish short and long survival periods. Additionally, we observed a significant discrepancy in tumor classification between expression layers. Overall, our integrative findings establish proteomic heterogeneity in GBM as a gateway to understanding poor patient survival.

## Introduction

Glioblastoma (GBM) is the most common high-grade adult brain tumor. Despite aggressive treatment combining radio- and chemotherapy, as well as gross-total resection of the tumor, disease usually progresses and the median survival time of patients is less than two years (Ostrom et al., 2015; Patel et al., 2019; Strobel et al., 2019; Stupp et al., 2017). A major challenge in GBM therapy is the tumor heterogeneity, which has been characterized on the genomic and transcriptomic levels.

Several studies conducted by the Cancer Genome Atlas (TCGA) described the genomic landscape of GBM tumors, defined by tumors invariably bearing EGFR amplification, TP53 and NF1 mutations in addition to other genetic aberrations (Cancer Genome Atlas Research Network, 2008). GBM classification based on gene expression signatures has established transcriptional heterogeneity (Nutt et al., 2003; Phillips et al., 2006). The analyses identified molecular subtypes associated with prognosis, which were later on refined based on RNA-sequencing (RNA-seq) (Verhaak et al., 2010) and more uniform sample selection (Wang et al., 2017) to yield three subtypes termed Proneural (PN), Mesenchymal (MES) and Classical (Cla). More recent studies, however, found little to no association between subtypes and prognosis (Wang et al., 2017). This is in part due to elimination of IDH1-mutant patients from the analyzed cohorts, as they are known to harbor less aggressive tumors and were previously defined as part of the PN subgroup (Wang et al., 2017). Beyond bulk tumor analysis, single-cell RNA-seq studies have shown that several transcriptional signatures exist within single tumors, representing different biological processes such as hypoxia and cell cycle (Patel et al., 2014). Furthermore, Neftel *et al*. (2019) recently showed that transcriptional heterogeneity of GBM tumors converges to four signatures that resemble mesenchymal and normal brain lineage stages and represent four tumor cell subpopulations (Neftel et al., 2019).

Omics-based studies have largely contributed to elucidation of these sources of heterogeneity. However, we hypothesized that tumor heterogeneity at the proteomic level should also be comprehensively evaluated, given that mass spectrometry (MS) based proteomics has become an integral part of cancer research, shedding light on the functional profile of the cancer cell (Coscia et al., 2018; Harel et al., 2019; Mertins et al., 2016; Pozniak et al., 2016; Tyanova et al., 2016a; Vasaikar et al., 2019; Yanovich et al., 2018; Zhang et al., 2016). In GBM, early proteomic studies have mostly utilized MALDI-TOF technology and applied it to find secreted tumor biomarkers from either cell lines or patient samples (Gautam et al., 2012; Kumar et al., 2010; Thirant et al., 2012). Other studies applied the technology to study the tumor cell proteome of GBM xenograft rat models (Rajcevic et al., 2009) or of cell lines undergoing different treatments (Puchades et al., 2007). More recent proteomic analyses of gliomas applied higher resolution MS techniques to study clinical samples of grade II-IV gliomas (Buser et al., 2019) and to identify proteomic differences between gliomas of various grades and genomic alterations (Djuric et al., 2019). However, GBM represents a small fraction of the tumors analyzed in these studies.

Integration of multiple omics-based methods further advances the comprehensive view of cancer. Specifically, proteogenomics combines MS-based proteomic data with whole exome and/or whole transcriptome sequencing in order to identify the functional outcome of genetic alterations and to evaluate the differences between expression layers. Recently published studies by the Clinical Proteomic Tumor Analysis Consortium (CPTAC) and others include proteogenomic analyses of ovarian, breast, colon, hepatocellular and gastric cancer (Gao et al., 2019; Mertins et al., 2016; Mun et al., 2019; Zhang et al., 2014; Zhang et al., 2016). Among other findings, these studies report the correlation between mRNA and protein levels of the same tumor tissues, and show a median correlation that is usually modest, ranging from 0.28 in gastric cancer to 0.54 in hepatocellular carcinoma. This further supports the potential benefit that can be provided by the proteomic layer.

In the current work, we present the first proteogenomic dataset of GBM clinical samples to date. We have assembled a cohort of 87 GBM patients of varying survival rates and performed MS-based proteomics analysis as well as RNA-seq in order to identify the molecular differences associated with survival and examine the contribution of each layer to GBM landscape. We show that the protein layer is more significantly associated with patient survival, but in addition, RNA-protein integration identifies clear patterns of layer-specific and layer-common processes specifically contributing to either short-term or long-term survival periods of patients. Furthermore, we compare our data to published single-cell RNA-seq of GBM tumors and evaluate the RNA-protein variability within single-cell based tumor subpopulations. We found that while all signatures of the four subpopulations tend to have high RNA-protein correlation, each signature is associated differently with survival. Altogether, these results highlight the potential of proteogenomics to further stratify heterogeneity in GBM tumors and identify processes contributing to poorer survival.

## Results

### Proteogenomic association with GBM clinical parameters

To examine the proteogenomic association with patient survival, we carefully selected samples to encompass a relatively large range of survival rates, from under three months to over ten years. We collected tumor specimens from 87 patients that were all pathologically defined as GBM, for which we generated either MS-based proteomics data, RNA-seq data or both (**Figure 1A**). After quality filtration, we continued the analysis with 84 samples out of which 54 had high-resolution proteomic data, 65 had RNA-seq data and 32 had both (**Figure 1B**). Using TMT-10plex chemical labeling, we identified 7096 proteins in total, out of which 4567 were used for downstream analyses (see Experimental Procedures). To account for stromal contamination of the transcriptomic samples, we filtered the RNA-seq gene list according to a bona-fide glioma (BFG) gene list generated by Wang et al. (2017), and performed the downstream analyses with the resulting 11,459 genes (see Experimental Procedures). To minimize sample variation, we took mostly IDH1-WT, untreated primary tumors. To initially increase the number of samples with IDH1 mutation annotation, we classified the samples using a published RNA-seq based signature (Baysan et al., 2012), and found eight samples to be IDH1-mutant, out of which only one had both RNA and proteomic data (**Figure 1C, Figure S1A, and Table S1**).

**Figure 1:**
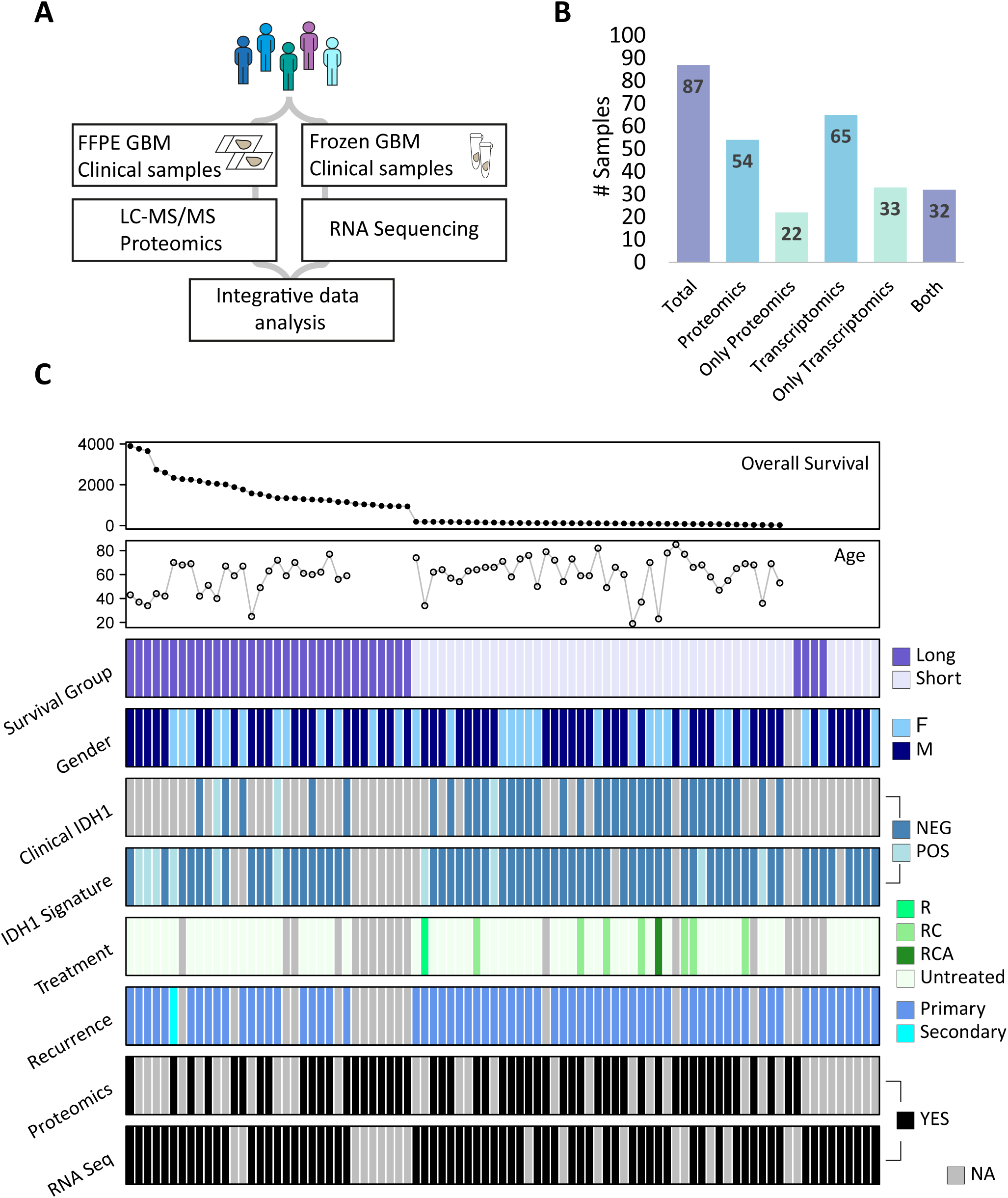
generation of proteogenomic cohort of glioblastoma patients (see also Figure S1). A) For the proteogenomic workflow, samples were collected from resected tumors and were subject to mass spectrometry analysis and RNA-sequencing. B) Number of samples in total and per data type. In each column, different color represents number of samples that have only the data type in the column specified. C) Heatmap describing samples collected from 87 GBM patients. Clinical parameters are indicated (R=Radiation therapy, RC=Radiation and chemotherapy, RCA=Radiation and chemotherapy combined with Avastin). Missing values in overall survival panel indicate precise survival days were not available.

Our main aim was to associate between RNA and protein profiles and the patient clinical parameters: survival, prior treatment, recurrence, gender and age. To find functionally related genes and their clinical associations, we performed weighted gene correlation network analysis (WGCNA) independently for transcriptomics and for proteomics, and used only primary IDH-WT tumors (n=52 proteomics, n=56 transcriptomics). We found 41 and 34 modules in proteomics and transcriptomics data, respectively. We then calculated the correlation between each module’s eigengene and clinical traits of interest and retained only significant (p<0.05) correlations. In both expression layers we identified modules that correlate significantly to age, gender and treatment. The major difference was observed in survival correlating modules: while six proteomic modules correlated with survival (three of them positively and three of them negatively), none of the RNA modules did (**Figure 2A**). It is worth noting that while some of these survival-associated modules correlated only with survival, the long-survival modules also presented a significant and opposite correlation to treatment. However, exclusion of treated samples resulted in similar long-survival correlating modules (**Figure S2A and Table S2**). It also yielded a single RNA survival module that contains 32 genes and was not enriched for any biological processes. Interestingly, when including IDH1-mut samples in the RNA WGCNA, we do find modules associated with survival. However, each of these modules is also significantly correlated to IDH1 status, indicating that transcriptomic expression profiles do not inform survival beyond IDH1 status (**Figure S2B**). To confirm the proteomic association with survival, we calculated the correlation with survival of each gene in each survival-correlating module (from Figure 2A; gene significance), and selected the top two proteins in each module. Kaplan-Meier analysis of these 12 proteins (two per each of the six survival-related modules) showed that they are all significantly associated with patient survival (**Figures 2B and 2C**). Interestingly, one of the long survival modules is represented by two immune proteins (CD5L and IGHM). Other than these, this module consists of multiple complement system components, implying a potential anti-tumorigenic role (for full details regarding module membership and functional enrichments see **Table S2**).

**Figure 2:**
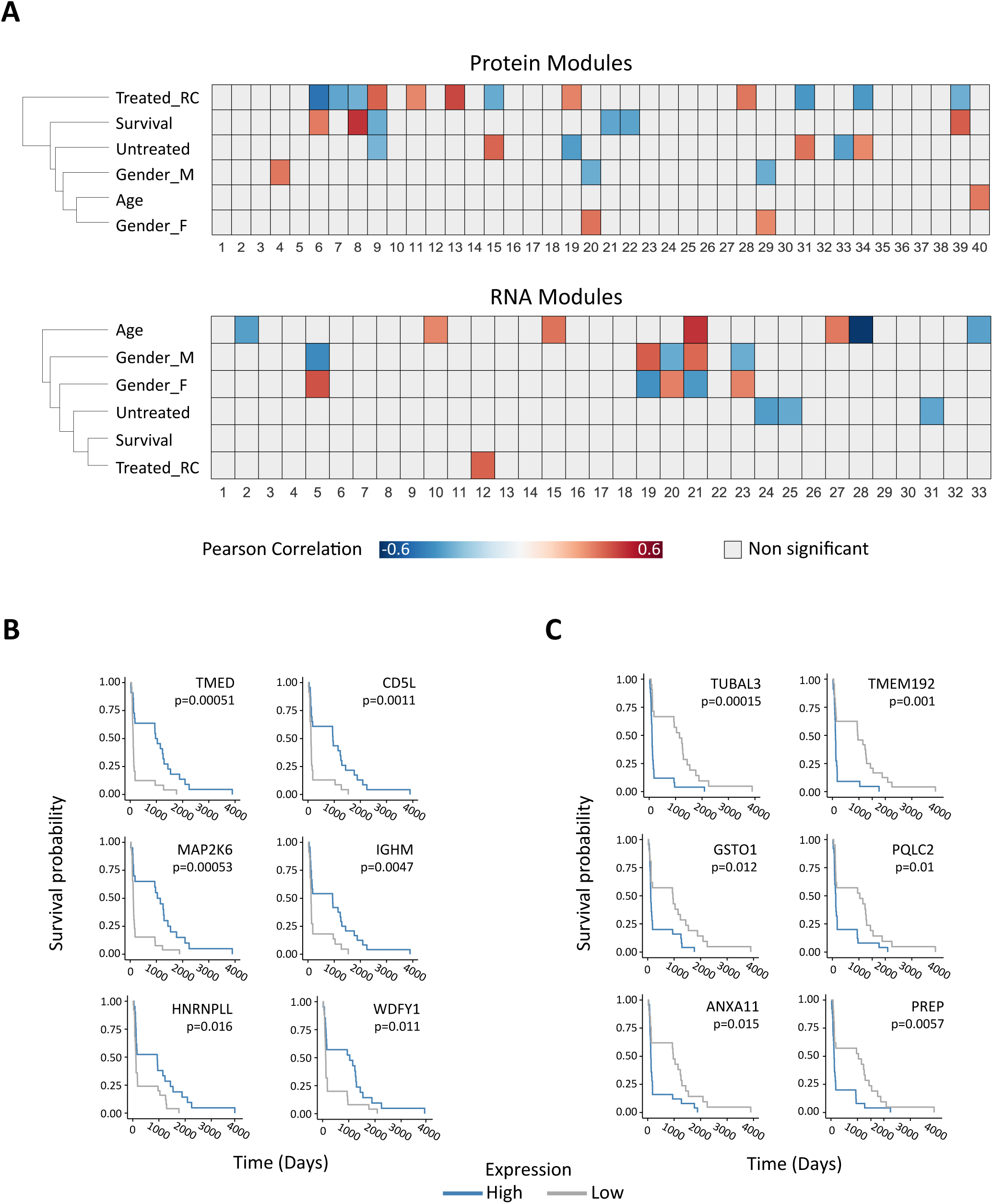
RNA and protein associations to clinical parameters (see also Figures S2A and S2B). A) Heatmap describing modules resulting from independent WGCNA analyses in protein (top) and RNA (bottom) (Treated_RC=treated with radiation and chemotherapy). Significant Pearson correlations (p<0.05) to clinical parameters are indicated in red to blue color, as indicated in the color bar below. Module 0 (grey module), which includes features below the module adjacency cutoff is not shown. B+C) Kaplan-Meier plots of selected proteins from short (B) and long (C) survival modules show significant association with survival (log rank p<0.05).

### Unsupervised clustering of samples differs between the proteomic and transcriptomic levels

In order to further compare the RNA and protein layers, we performed unsupervised classification independently on the proteomic data (54 samples) and transcriptomic data (65 samples) using consensus clustering algorithm (Monti et al., 2003). In a separate analysis, we classified the tumors in our cohort according to established RNA-based signatures (Wang et al., 2017). Out of 65 analyzed RNA samples, 19 were defined as the classical (Cla) subtype, 16 as mesenchymal (MES) and 17 as proneural (PN, hypergeometric p-value < 0.05, see Experimental Procedures). The transcriptomic consensus clustering resulted in three robust clusters that match the transcriptional subtypes (group 1 is Cla, group 2 is PN and group 3 is MES) (**Figure 3A and Figure S2C**). The proteomic classification also resulted in three groups (**Figure 3B and Figure S2D**), however with only 25% of the overlapping samples clustering similarly in both layers. In order to try and match between the groups identified in each layer, we looked for differentially expressed genes or proteins in each classification (ANOVA test FDR<0.01), and examined their enriched functionalities. Hierarchical clustering showed that the differentially expressed features in each layer divide into three clusters. The functional enrichment in the RNA clusters mostly recapitulated the known gene expression signatures for each of the subtypes (Verhaak et al., 2010) (Fisher’s exact test, FDR<0.02; **Figure 3C**). Two clusters in each classification have distinct biological processes, such as NF-ᴋB signaling and epigenetic regulation in RNA clusters, or proteasomal regulation and translation in protein clusters (**Figures 3C and 3D**). Intriguingly, one cluster was functionally similar in RNA and protein, displaying a neuronal profile and enriched for processes such as synaptic transmission and neuron generation. This cluster corresponds to protein group 3, and to RNA group 2, which consists of mostly PN samples. However, only two of the shared samples overlapped between these two groups, even though protein networks associated with these enrichments include overlapping and inter-connected members, such as guanine nucleotide-binding proteins (GNGs) and the neuronal calcium sensor NCALD. Beyond these, each cluster has layer-specific members, such as several subunits of the glutamate receptor (GRIK genes) in the RNA network and protein transport-associated proteins of the AP2 complex in the protein network (**Figure 3E**). These results show that neuronal features appear in both layers, but are apparent in a different set of tumors. This discrepancy may partially result from internal tumor heterogeneity. Nevertheless, it highlights proteomics as another layer of tumor heterogeneity in GBM (see **Tables S3A-S3D** for lists of differentiating features and significantly enriched processes). Given the profound differences in the clustering analyses, we examined the global RNA-protein agreement of processes, and specifically the processes observed in the classification analysis. To that end, we combined the two datasets and calculated the Spearman rank correlation between RNA and protein expression over 4,514 genes quantified in both layers of the 32 shared samples. The median Spearman correlation was rather modest (r=0.16 for 67% positive correlations, **Figure 3F**), compared to published proteogenomic studies (Mertins et al., 2016; Zhang et al., 2016), and we assume that the lower correlation results from the different tumor blocks used for each analysis. Nevertheless, the biological processes enriched in each extreme of the RNA-protein agreement axis (1D annotation enrichment test, Benjamini-Hochberg FDR<0.05) recapitulate known findings regarding shared and layer-specific processes (Mertins et al., 2016; Zhang et al., 2014; Zhang et al., 2016). For example, ribosome and translation processes are enriched within the negatively correlating genes, which may explain the identification of these processes only in the proteomic classification (**Figure 3D**). Processes such as proteasome and extracellular matrix appear to be enriched with positively correlating genes (**Figures 3G and 3H, Table S3E**).

**Figure 3:**
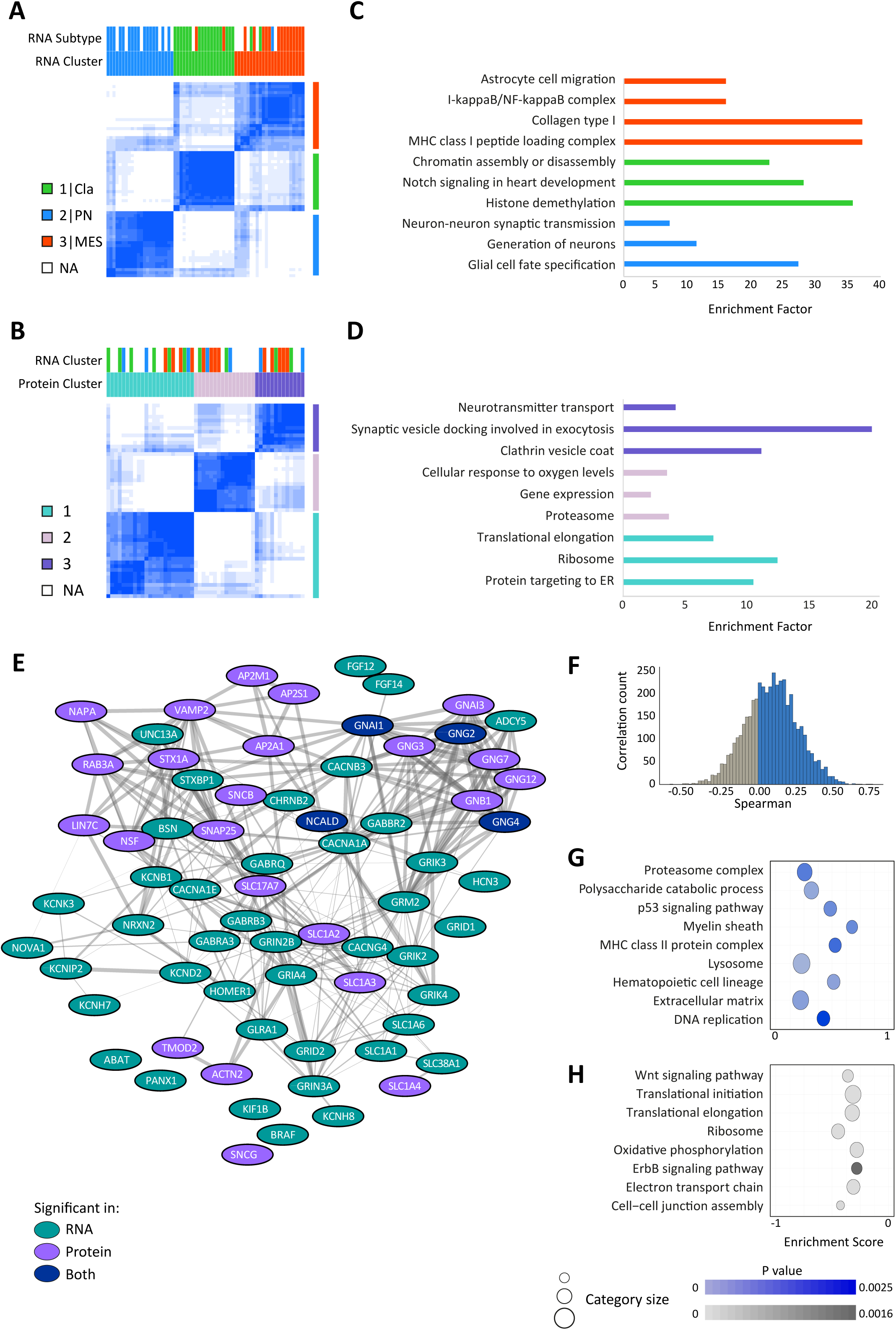
comparing unsupervised clustering of each expression layer (see also Figures S2C and S2D). A+B) Consensus clustering heatmap showing three clusters found in either transcriptomic (A) or proteomic (B) data. Blue color represents sample consensus score. Cluster annotations are indicated. C+D) Enriched biological processes and pathways in each cluster in each classification (Fisher exact test, FDR < 0.02). E) Protein-protein interaction network of the “synaptic transmission” category in protein and RNA. Edge width represents level of confidence for the interaction evidence. Color represents layer (purple=protein, turquoise=RNA, dark blue = both). F) Global RNA-protein correlation based on 32 overlapping samples. G+H) Enriched biological processes within positively- (G) or negatively- (H) correlating genes. Circle size stands for GO category size and p-values are indicated by respective colorbars.

### Integrating RNA and protein to identify layer-specific contribution to survival

The global correlation analysis as well as the layer-specific classification show that the differences between RNA and protein potentially represent yet another layer of inter-tumor heterogeneity in GBM, in the gene expression level. To evaluate the contribution of each layer to survival, we filtered the data to retain only genes whose expression was quantified in both layers (n=3407) and then calculated the correlation between patient survival, and the gene’s protein or RNA expression across all patients. Next, using the correlation coefficients, we clustered the significantly correlating genes in either layer (permutation based adjusted p-value<0.1, **Figure 4A**). For each of the resulting seven clusters, we checked whether it was enriched for protein-significant correlations, RNA-significant correlations or both (Fisher exact test Benjamini-Hochberg FDR<0.02), and whether it is associated with short or long-term survival (**Figure 4B**). We termed the clusters according to their layer enrichment and to their association with long or short survival: while some genes correlated highly and negatively with survival in both RNA and protein (common-short), some correlated highly and positively in both layers (common-long). Interestingly, the latter is the smallest cluster, indicating that RNA and protein correlate similarly to survival mostly when survival is shorter, and suggesting that longer survival is defined by layer-specific processes. Furthermore, several clusters contain genes correlating to survival only in one layer, and one cluster was enriched for genes whose survival-related behavior was altogether opposite in the two layers. Controlling for IDH1 mutation status produced similar results except for several common correlations and processes (**Figure S3**).

**Figure 4:**
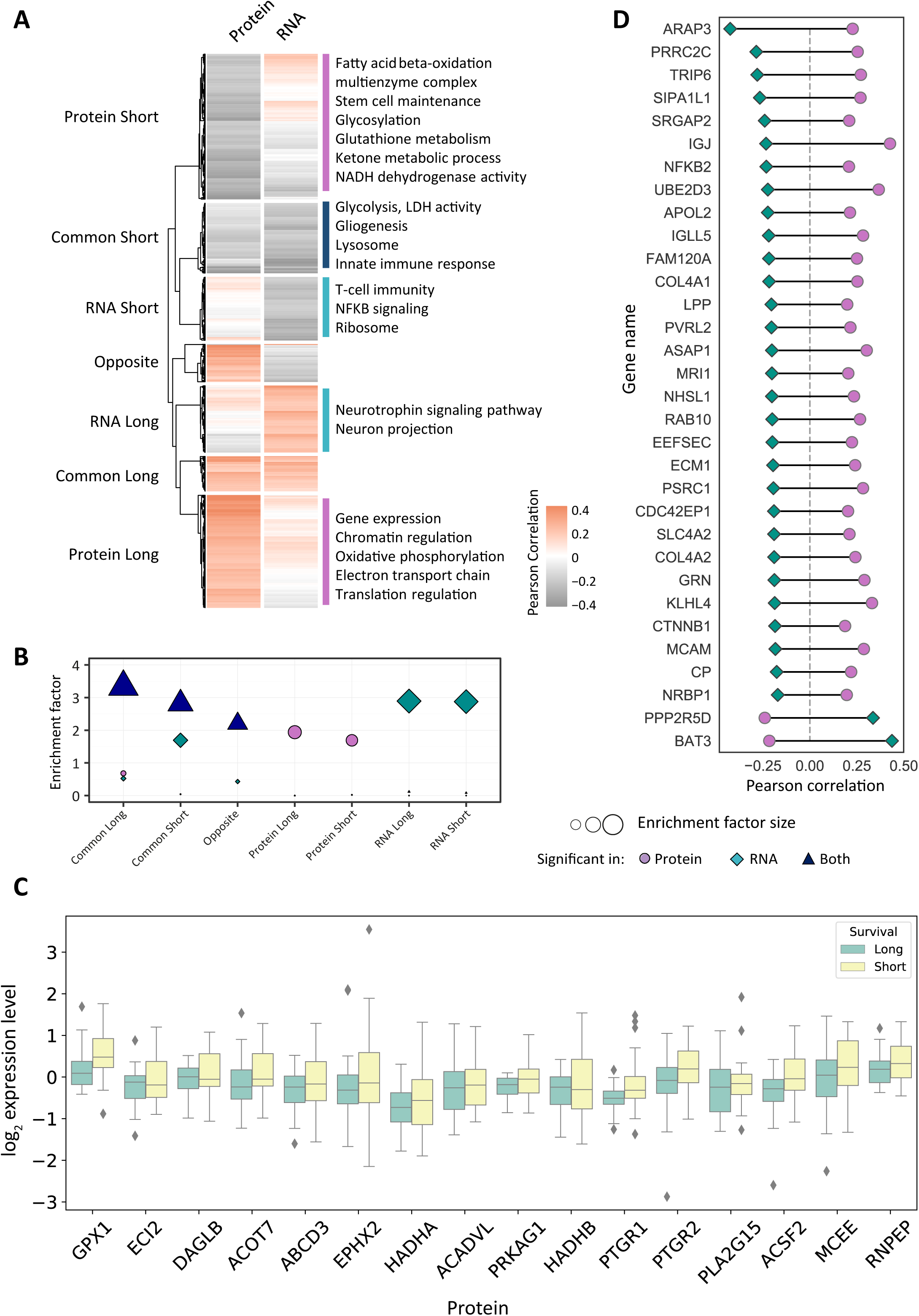
Integrated analysis of survival (see also Figure S3). A) Hierarchical clustering of correlation coefficients of 1320 genes significantly correlating to survival in either RNA or protein (adjusted p-value<0.1) identifies seven clusters. B) Each of the seven identified clusters is enriched for layer-specific significant correlations. Common and opposite clusters are enriched for genes significantly correlating to survival in both RNA and protein. Shape type indicates expression layer and shape size corresponds to enrichment factor. C) Boxplots of proteins taken from the “Protein-short” pattern and belong to the Beta-oxidation GO category. We divided the samples to two groups based on survival (short survival = less than 6 months, long survival = more than 1.5 years). D) 32 genes of the “opposite” cluster that have opposite and significant correlations to survival, and represent mostly cell-ECM interaction.

Gene ontology (GO) enrichment analysis showed that enriched processes in the common-short cluster include glycolytic metabolism, inflammatory response and lysosomal activity. These processes recapitulate known biological aspects of GBM tumor aggressiveness (Murat et al., 2009; Reynes et al., 2011; Stettner et al., 2005; Wolf et al., 2011; Yeung et al., 2013). We did not find enriched processes in the common-long cluster, presumably due to its small size.

Long-survival processes in the RNA level include neurotrophin signaling pathway and neuron projection, and the genes belonging to these processes are MAPK signaling related genes such as KRAS, MAPK8, MAP2 and CRKL. MAPK8 is known for its role in neuronal development (Chang et al., 2003; Westerlund et al., 2011), as well as MAP2.

The protein-long cluster, in addition to gene expression and chromatin regulation processes, is enriched for oxidative phosphorylation and the electron transport chain. This could be interpreted as a metabolic mirror image of the short-term survival glycolytic metabolism, suggesting an attenuated Warburg effect in less aggressive GBM tumors.

Layer specific processes are also found in association with short-term survival. The RNA-short cluster is enriched for T-cell immunity and NF-ᴋB signaling, and thus further supports the immune activation observed in the common-short cluster. The protein-short cluster is enriched for stem cell maintenance, known to be a marker for poorer prognosis in GBM. It also reveals a metabolic profile that goes beyond glycolysis, and includes β-oxidation and ketone body metabolism (**Figure 4C**, see **Table S4** for all significantly enriched processes). Despite relying primarily on glucose metabolism, fatty acid oxidation also has a potential role in glioma cell growth (Lin et al., 2017, see Discussion).

Interestingly, we found a cluster of 101 genes that correlate oppositely to survival. This cluster includes a group of genes related to cell-extracellular matrix interaction (ECM1, COL4A1, NID2 and ITGA1), and the NF-ᴋB2-RELA transcription factor complex. Out of the 101 genes, 32 have an opposite and significant correlation in both RNA and protein (**Figure 4D**). Among these genes, we found the Wnt pathway member β-catenin (CTNNB1), which is known to be primarily regulated by protein degradation. β-catenin is mostly located in the cytoplasm or membrane in GBM, and rarely in the nucleus (Denysenko et al., 2016; Liu et al., 2011; Yano et al., 2000), which may explain its association with longer survival. Another interesting gene is the melanoma cell adhesion molecule MCAM (CD146), which was found to be highly expressed in glioma stem cells (Yawata et al., 2019). However, its role in GBM is yet to be fully understood.

### Validating survival related processes by integrating single-cell RNA data

Given the reported intra-tumor heterogeneity in GBM, and the single cell populations identified by single cell RNA-seq (Neftel et al. Cell 2019), we next examined whether integration with the bulk proteogenomic data can add the clinical relevance to the identification of tumor cell subpopulations. Neftel *et al*. identified four different tumor subpopulations: mesenchymal (MES), astrocytic (AC), neural progenitor cell-like (NPC) and oligodendrocytic precursor cell-like (OPC) that are shared between patients, but have different relative frequencies in each tumor. For each subpopulation, they established a gene signature that enables calculation of the population fraction in bulk tumor analyses, and to further associate these with clinical features. Applying these signatures to our proteogenomic data, we examined whether different subpopulations could be associated with long or short-term survival. We ranked all the genes/proteins in our dataset according to the RNA/protein correlation with survival, and examined the enrichment of each of the four signatures in the positively or negatively correlating genes with survival (see Experimental Procedures). Analysis of the bulk RNA data showed significant association of AC and MES signature genes with shorter survival, and of OPC and NPC signature genes with longer survival (**Figure 5A**). Only the MES signature was also significantly enriched in the protein data, and lower significance of the protein level is expected given that the single-cell signatures were based on RNA analyses.

**Figure 5:**
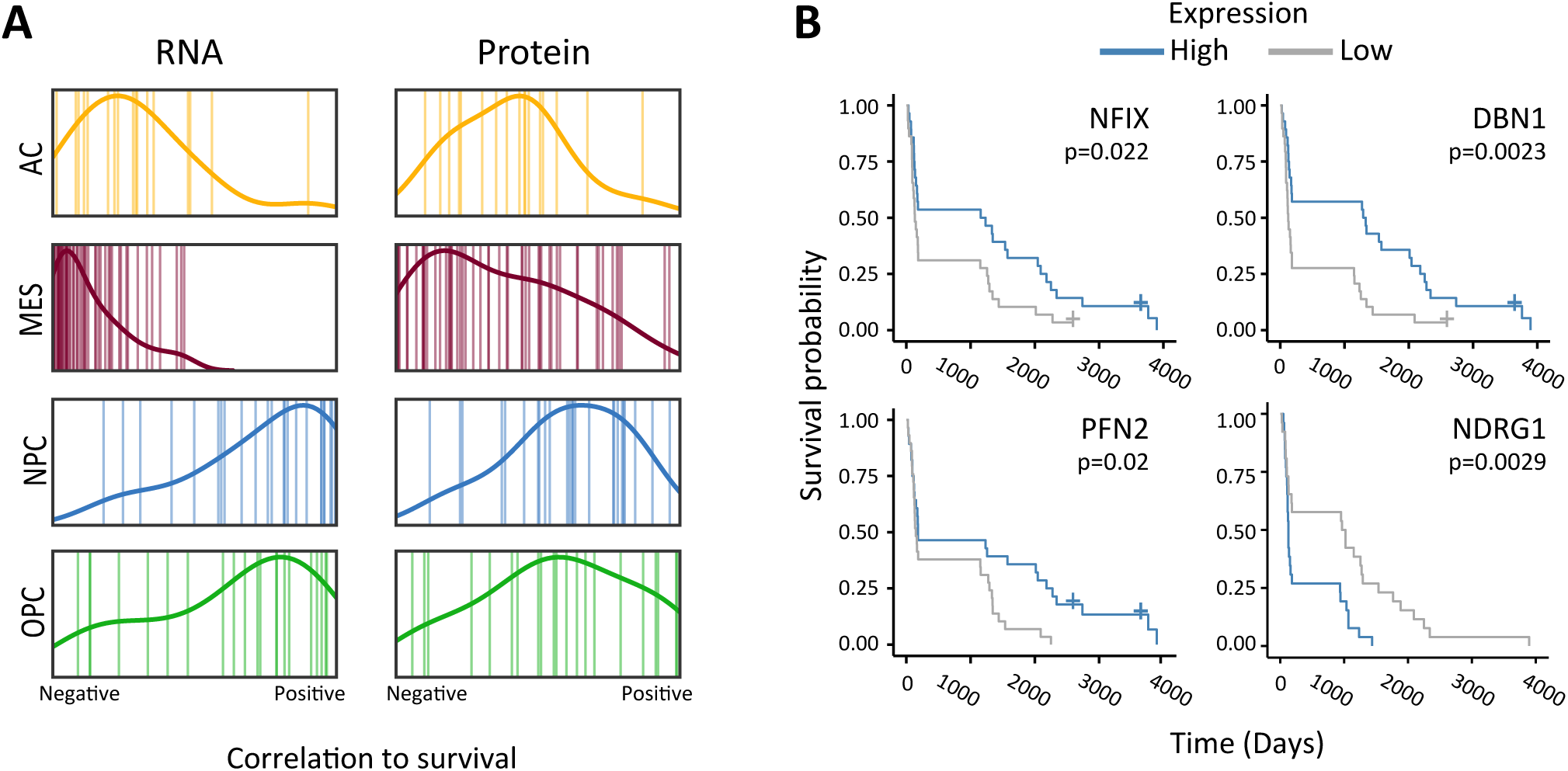
Integrating single-cell RNA-seq subpopulations and survival. A) Barcode plot showing the rank of each single-cell subpopulation signature in a scale of correlation to survival in either protein (right) or RNA (left). High rank indicates positive correlation to survival, while low rank indicates negative correlation. AC-astrocytic, MES-mesenchymal, NPC-neural progenitor cell-like, OPC-oligodendrocytic precursor cell-like. B) Kaplan-Meier plots of signature genes found to be significantly associated with survival in either RNA (NFIX, PFN2, DBN1) or protein (NDRG1).

Next, we examined whether individual population-specific genes can serve as survival markers in each expression layer. For the longer survival subpopulations (NPC and OPC), we took the top five ranks (positive correlations), and for the shorter survival subpopulations (AC and MES), we took the lowest five ranks (negative correlations). This formed a list of 40 genes, out of which seven genes were common to RNA and protein (**Table S5**). Kaplan-Meier survival analysis found four genes to be significantly associated with survival (log-rank test p-value < 0.05): three in the RNA level (DBN1, PFN2 and NFIX), and one in the protein level (NDRG1) (**Figure 5B**). Altogether, these results enabled us to link between cell subpopulation and survival, and identify potential prognostic markers.

## Discussion

In this work, we investigated the proteomic contribution to GBM tumor heterogeneity, and its association with patient survival. When performing the two “omic” WGCNA analyses separately, we found proteomic expression profiles that correlate with survival in IDH1-WT tumors, but could not find such profiles in RNA data. This finding recapitulates other studies that utilized RNA-seq on IDH1-WT tumors and also did not observe a clear association with prognosis (Wang et al., 2017). However, when we applied an integrative approach that combined protein expression, RNA expression and survival correlation, we were able to identify survival patterns in both layers, and define whether they are shared between expression layers or layer-specific. Since our proteomic cohort included only one IDH1-mut sample, follow-up studies could refine this analysis by comparing the IDH1-WT and mutant proteomic profiles of GBM, as recent evidence suggests a prominent proteomic difference between these groups in lower grade gliomas (Djuric et al., 2019).

Our analyses highlight three main mechanisms to be associated with survival: Immune processes, metabolic processes and developmental processes. We found short survival to be associated with inflammation and glycolytic metabolism, both with established roles in tumor growth and aggressiveness in GBM (Reynes et al., 2011; Waters et al., 2019; Yeung et al., 2013). Interestingly, while NF-ᴋB signaling and T cell immunity negatively correlate with survival and reflect a pro-tumorigenic inflammatory response, the WGCNA results show a positive association between immunity and survival in the form of immunoglobulins and complement system components, suggesting a potential anti-tumorigenic immune response. Furthermore, the proteomic layer provides an extended metabolic context for patient survival; linking increased fatty acid oxidation with short survival, and oxidative phosphorylation (OXPHOS) with longer survival time. These findings are supported by the identification of the role of fatty acid oxidation in tumor growth and oxidative stress mitigation (Duman et al., 2019; Pike et al., 2011) and the role of OXPHOS in tumor suppression and mitochondria-promoted apoptosis in GBM (Michelakis et al., 2010). Additionally, our integrative analysis revealed that stemness-development axis is also evident in the protein level. Traditionally, cancer stem cell populations in gliomas have been shown to possess higher tumorigenic capacity, and as such were considered a promising therapeutic target (Berger et al., 2004; Galli et al., 2004; Suva et al., 2014). However, while the association between stemness and aggressiveness was observed in lower grade gliomas (Tirosh et al., 2016; Venteicher et al., 2017), it is not straightforward in GBM tumors, in which multiple cells invariably express stemness markers (Patel et al., 2014). In addition, the high plasticity of GBM cells enables differentiated tumor cells to undergo de-differentiation, regulated by the tumor microenvironment, epigenetics and other factors (Dirkse et al., 2019). For example, it was recently shown that differentiation-inducing treatment in GBM cell lines is mediated by elevated mitochondrial metabolism and specifically OXPHOS (Xing et al., 2017). Our findings reinforce the functional connection between stemness, metabolism and overall survival in GBM. This suggests that further investigation of the proteomic profiles associated with stemness in shorter and longer survival might benefit stemness-targeted clinical efforts.

Internal tumor heterogeneity is an inherent limitation of omic data generated from bulk tissues. Indeed we performed some of the analyses by directly comparing the data types, however performing most analyses in a layer-independent examination and then comparing the results enabled us to overcome the partial overlap between RNA and protein samples. To address the constraint of samples not taken from the exact same tumor region, we further incorporated publically available single cell RNA-seq data. Our analyses show a similar trend for both expression layers (though as expected, significant in the RNA level only), wherein two single-cell subpopulations indicate short survival while two other subpopulations indicate longer survival. In order to substantiate the connection between RNA levels, internal tumor heterogeneity and patient outcome, it would be necessary to check whether the ratio between long and short subpopulations within a tumor reflects patient outcome, once single cell techniques enable the collection of larger patient cohorts. Furthermore, single cell proteomic analyses are expected to unravel whether the bulk inter-layer differences are also apparent in the individual cell populations.

The association mentioned above was only evident in the signature level. Effectively, only four genes showed a significant association with survival. One of these genes is NDRG1 (N-myc downstream regulated gene 1), which is a member of the mesenchymal signature. NDRG1 is a stress protein known to be induced in hypoxic conditions (Cangul, 2004) and to be downregulated in several cancer types while upregulated in others (Melotte et al., 2010). In brain tumors, while evidence was found for both good and bad prognostic effects (Sun et al., 2009; Weiler et al., 2014), it appears that NDRG1 is increased in GBM when compared to lower grade gliomas similarly in the RNA and protein levels (Said et al., 2009; Weiler et al., 2014), and that its upregulation confers chemotherapy resistance (Weiler et al., 2014). Our data support the role of NDRG1 as a poor survival marker, and propose it as a potential therapeutic target.

The discrepancy between RNA and protein was largest in the tumor classification analyses. While some of these differences may be apparent due to the internal tumor heterogeneity, previous classifications of other tumor types (e.g. breast cancer (Mertins et al., 2016; Yanovich et al., 2018)) did not show such major differences, despite analyzing distinct regions/tumors. In contrast, in GBM we found minor concordance between the functionalities of the RNA and protein clusters.

In conclusion, we presented here the first MS-based global characterization of protein expression in clinical samples of glioblastoma. Our proteogenomic analysis highlights the contribution of proteomic investigation to understanding cancer biology and association with patient survival. The identified proteomic and transcriptomic patterns shed light on the intriguing molecular heterogeneity of GBM tumors, by revealing novel functionalities related to patient survival and disentangling the contribution of each expression layer.

## Data Availability

The mass spectrometry proteomics data have been deposited to the ProteomeXchange Consortium via the PRIDE partner repository with the dataset identifier PXD018024.
The RNA-seq data have been deposited in NCBI's Gene Expression Omnibus (Edgar et al., 2002) and are accessible through GEO Series accession number GSE149009 (https://www.ncbi.nlm.nih.gov/geo/query/acc.cgi?acc=GSE149009).

## Acknowledgments

The Geiger laboratory has received partial Funding from the European Research Council (ERC) starting grant [639534], the Israel Science Foundation (grant 748/16) and the Gail White and Anne and William Cohen Multidisciplinary Brain Cancer Research Program. The Satchi-Fainaro laboratory has received partial funding from the European Research Council (ERC) Consolidator Grant Agreement no. [617445]-PolyDorm and ERC Advanced Grant Agreement no. [835227]-3DBrainStrom; The Israel Science Foundation (Grant nos. 918/14 and 1969/18); Israel Cancer Research Fund; Nancy and Peter Brown friends of The Israel Cancer Association (ICA) USA, in memory of Kenny and Michael Adler (20150909); and from the Morris Kahn Foundation. E.Y. is a fellow at the Gail White and Anne and William Cohen Multidisciplinary Brain Cancer Research program. P.O. thanks the Naomi Foundation for the Global Research and Training Fellowship in Medical and Life Sciences.

We thank Naama Knafo, Michal Harel and Lir Beck for their critical evaluation of the manuscript, and all members of the Geiger laboratory for fruitful discussions. We thank Tom Rabinowitz for technical assistance in cluster-based analyses.

RNA-seq was performed by Otogentics Coproration (Atlanta, GA, USA).

## Author Contributions

Conceptualization, G.Y.A., T.G. and R.S.F.; Formal Analysis and Investigation, G.Y.A.; Resources, R.G., R.S.F., P.O., E.Y., G.Y.A., A.D. and N.S.; Writing – Original Draft, G.Y.A. and T.G.; Writing – Review & Editing, G.Y.A., P.O., E.Y., R.G., N.S. and T.G.; Funding Acquisition, R.S.F. and T.G.; Supervision, T.G.

## Experimental Procedures

### Clinical samples acquisition

Frozen tissue blocks and formalin-fixed paraffin-embedded (FFPE) tissues were obtained from the Neurosurgery and Pathology departments of Tel Aviv Sourasky Medical Center, Tel Aviv, Israel. We acquired tumor samples from 87 patients, all taken from tumor resection surgery; 69 were pathologically defined as primary glioblastoma (GBM), one sample was secondary GBM and for 17 patients recurrence information was not available. Only eight patients were given treatment (radiotherapy and chemotherapy) prior to surgery. For two patients (L18 and L19), we had samples from two different foci. The cohort included tumors from 29 females, 49 males and 9 without gender information, with age range of 19 to 85 (median 62). Additionally, in order to perform a molecular analysis of patient survival, samples were specifically selected to have varying survival rates, ranging from less than three months to over 10 years (median ~5 months). All samples were obtained upon ethical approval from the IRB committee of the Tel Aviv Sourasky Medical Center.

### RNA extraction and sequencing

RNA was extracted from fresh frozen GB tissues of human patients using EZ RNAII isolation kit (Biological Industries, Bet Haemek, Israel) according to manufacturer’s instructions. Samples were homogenized in Denaturing Solution (0.5ml/50–100mg tissue) using GentleMACS homogenizer (Miltenyi Biotech, USA) program RNA-02. Homogenates were stored for 5 minutes at room temperature, then 0.4ml Water-saturated phenol was added followed by 0.09ml 1-Bromo-3-chloropropane (BCP) and vigorous shaking for 15 seconds. The resulting mixture was stored at room temperature for 10 minutes and then centrifuged at 12,000g for 15 minutes at 4°C. The aqueous colorless (upper) phase was transferred to a fresh tube followed by addition of 0.5ml isopropanol. Mixture was stored for 30 minutes at −20°C and then centrifuged at 12,000g for 8 minutes at 4°C. Supernatant was removed and RNA pellet was washed (by vortexing) with 1ml 75% ethanol, then centrifuged at 7,500g for 5 minutes at 4°C. Ethanol was removed, RNA pellet was air-dried for 20–30 minutes and then dissolved in 100μl of DNAse RNAse-free water (Biological Industries, Bet Haemek, Israel) by incubating for 10–15 minutes at 55°C. RNA samples were kept at −80°C until sequencing. RNA Integrity Number (RIN) was determined for each sample using the 2200 TapeStation system (Agilent, CA, USA). RNA libraries were prepared according to Illumina protocols. Paired-end RNA sequencing data (read length 100 base pairs, designated 20 million reads per sample) was generated on the Illumina HiSeq 2500 at Otogenetics Corporation, Atlanta, GA USA.

### Protein extraction

54 FFPE blocks were macro-dissected from tissue slices by overlaying H&E staining, in order to enrich for cellular areas and exclude stromal components. Dissected samples were lysed in 50% 2–2–2 trifluoroethanol (TFE) in 25mM ammonium bi-carbonate (ABC), incubated with 5mM Dithiothreitol (DTT) and alkylated with 15mM Iodoacetamide (IAA). Prior to protein digestion, samples were diluted 1:10 with 5mM ABC, and then digested overnight with LysC/Trp mix and Trypsin in an enzyme to protein ratio of 1:100 and 1:50, respectively. Prior to labeling, clean-up of digested peptides was performed using C18 Stage-Tips. We proceeded to tandem-mass-tags (TMT) 10plex labeling according to the manufacturer’s instructions (Pierce). The 54 samples were divided into six sets of 10plex-TMT, while the tenth sample in each set consisted of a tumor-mix to be used as a quantification standard between different sets. Following labeling, samples were combined and vacuum-concentrated, and then resuspended in 0.1% trifluoroacetic acid (TFA). Resuspended samples were loaded onto high-pH reverse phase columns (Thermo Scientific) for sample fractionation. Each 10plex-TMT set was fractionated into eight fractions according to the manufacturer’s instructions. Resulting fractions were then vacuum-concentrated and resuspended in MS loading buffer (2% acetonitrile, 0.1% formic acid).

### Liquid Chromatography – Mass Spectrometry (LC-MS) Analysis

Peptides were separated in the Easy-nLC 1000 nano-HPLC system (Thermo Fisher Scientific) using reverse phase chromatography on a C18 Easy-Spray column; and loaded to the Q-Exactive HF mass spectrometer (Thermo Fisher Scientific). Each sample ran for a 128-min gradient of water and 80% acetonitrile, with an MS resolution of 120,000 (scan range 350–1400 m/z, ion target value of 3e6 and maximum injection time of 100 ms) and MS/MS resolution of 60,000 (scan range 200–2000 m/z, ion target value of 1e5 and maximum injection time of 60 ms). In every MS scan, the top 15 most abundant peaks were selected for higher-energy collision dissociation (HCD) fragmentation.

### Protein Identification and Quantification

MS raw files were analyzed using MaxQuant software version 1.6.2.6 (Cox and Mann, 2008; Tyanova et al., 2016b). Peptide search was performed using the Andromeda search engine (Cox et al., 2011) against the Uniprot human protein database release April 2018, with 1% false discovery rate (FDR) at the PSM and protein levels. MS level mass tolerance was set to 4.5 ppm. Peptides were allowed to have methionine oxidation and N-terminal acetylation as variable modifications and cysteine carbimdomethyl as a fixed modification. Quantification was defined based on MS2 reporter ion intensity with TMT channels correction factors supplied by the manufacturer.

### Multiomic data analysis

#### RNA-sequencing data acquisition and analysis

Resulting reads in FASTQ format were trimmed and quality filtered using Trimmomatic (Bolger et al., 2014). We used Salmon quasi-mapping tool (Patro et al., 2017) for expression quantification against the transcriptome compiled from the Ensembl human hg38 genome assembly. We used the R package DESeq2 (Love et al., 2014) to convert transcripts per million (TPM) quantifications to gene level, as well as to perform log_2_ transformation and variance stabilizing transformations (VST) normalization of the counts for downstream analysis. Three samples were removed from downstream analysis due to low alignment rates. To account for stromal contamination of the samples, we filtered the RNA-Seq gene list according the bona-fide glioma (BFG) gene list generated by Wang et al. (2017), and performed all downstream analyses (except for the global protein-RNA correlation) with the resulting 11,459 genes.

#### Proteomic data pre-processing and statistics

We identified 7096 proteins in total in 54 patient samples and 6 control channels (5422 per sample on average). In order to retain high-quality quantifications, we filtered the data to contain only proteins that were quantified in all six standard channels. This resulted in 4567 proteins, for which missing value imputation was performed sample-wise by drawing values from a normal distribution with a width of 1.5 and down shift of 0.5 standard deviations of the specific sample. Unless otherwise specified, downstream analyses were then performed on calculated protein ratio between each sample and its corresponding standard. As samples of different survival rates were evenly distributed between TMT sets, we performed linear modelling to eliminate small TMT batch effects using R limma package (Ritchie et al., 2015). (**Figure S1B**).

#### Protein-RNA correlation

Out of a total of 84 samples, 32 had both proteomic and transcriptomic data, while 20 had only proteomic data and 32 had only RNA data. To calculate the protein-RNA correlation we used only the 32 samples for which we had data in both layers. We then matched between proteomic and transcriptomic data based on gene names and calculated the Spearman rank correlation coefficient between gene and protein expression for 4514 genes. To calculate the biological enrichments of either negatively or positively correlating genes we used 1D annotation analysis in which genes are ranked in ascending order according to their correlation, and genes of each biological category are tested for having significantly high or low ranks, as described (Cox and Mann, 2012).

#### Weighted gene correlation network analysis (WGCNA) and unsupervised clustering

WGCNA was performed using the WGCNA R package (Langfelder and Horvath, 2008) and WGCNA implementation in Perseus software (Rudolph and Cox, 2019). We used a soft-threshold beta power=16 to create a robust signed network. Network was then clustered using the algorithm default parameters. We used Pearson correlation to calculate the correlation between the module eigengene and clinical annotations.

For unsupervised classification of the RNA and the proteomics data we used consensus clustering algorithm implemented in ConsensusClusterPlus R package (Monti et al., 2003; Wilkerson and Hayes, 2010), with subsampling of 80% of the samples and 10 as maximum k (number of clusters). Before classification, samples were z-score normalized followed by protein/gene expression z-score normalization. Clustering results were evaluated visually in the resulting consensus matrix (one matrix per each k) as well as quantitatively using the cumulative distribution function of the area under the curve for each k (**Figures S2C and S2D**).

#### Integrated Pattern analysis

For the integration of proteomic and transcriptomic datasets we matched the two matrices based on gene names, filtered out genes that were not quantified at all in each one of the two datasets (n=3407) and merged genes based on gene name (n=3354). For the correlation analysis, we only kept the samples for which we had survival information (n=52 in protein, n=56 in RNA) and calculated Pearson correlation twice for each gene: between protein expression and survival, and between gene expression and survival. We also calculated a permutation based p-value for each correlation, by scrambling the expression data within samples and repeating the procedure 1000 times. For downstream analysis we kept genes with a significant correlation to survival (permutation based adjusted p-value<0.1) for either RNA or protein. The resulting 1426 genes, represented by their correlation to survival in each layer, were then hierarchically clustered. Each cluster was then evaluated based on the correlation pattern of its genes. Fisher enrichment test (FDR=0.02) tested whether each cluster was enriched for significant survival-protein correlations or survival-RNA correlations. Together with the directionality of the correlation, we were able to name each cluster as either “protein”, “RNA” or “both”; and as associated with either long-term or short-term survival.

#### Integration of published single cell data

Single cell RNA signatures were downloaded from Neftel *et al*. (2019). We determined the dominant single cell-based subpopulation as described (Neftel et al., 2019). Briefly, we calculated the average expression of each subpopulation’s signature genes. We compared it to background expression, created by randomly drawing 100 genes from the gene’s expression bin, for each gene in the signature. This resulted in four scores for each sample, one for each subpopulation. The subpopulation that received the highest score was considered as most dominant in that sample. Before this calculation, the genes in each signature were filtered to adjust for bulk tumor analysis, as described (Neftel et al., 2019). Kaplan-Meier analysis and logrank test were performed using R’s survival and survminer (https://cran.r-project.org/web/packages/survminer/index.html) packages.

All analyses were performed using the Perseus software (Tyanova et al., 2016c), R and Python. Biological annotations were taken from Gene Ontology (GO) and Kyoto Encyclopedia of Genes and Genomes (KEGG).

## Data availability

The mass spectrometry proteomics data have been deposited to the ProteomeXchange Consortium via the PRIDE partner repository with the dataset identifier PXD018024. The RNA-seq data have been deposited in NCBI’s Gene Expression Omnibus (Edgar et al., 2002) and are accessible through GEO Series accession number GSE149009 (https://www.ncbi.nlm.nih.gov/geo/query/acc.cgi?acc=GSE149009).

## Supplementary Tables

**Supplementary Table S1:** cohort details and clinical information.

**Supplementary Table S2:**module membership and functional enrichments of WGCNA

- Table S2A: module membership of each WGCNA analysis
- Table S2B: Functional enrichments in protein modules with significant correlations (Fisher Exact test, FDR<0.02)
- Table S2C: Functional enrichments in RNA modules correlating to clinical traits (Fisher Exact test, FDR<0.02)

**Supplementary Table S3:**significant features and functional enrichments within RNA and protein consensus clusters and RNA-protein correlation

- Table S3A: significantly differentiating proteins (protein consensus clusters, ANOVA test FDR<0.01)
- Table S3B: significantly differentiating genes (RNA consensus clusters, ANOVA test FDR<0.01)
- Table S3C: Functional enrichments in protein consensus clusters (Fisher Exact test, FDR<0.02)
- Table S3D: Functional enrichments in RNA consensus clusters (Fisher Exact test, FDR<0.02)
- Table S3E: Functional enrichments within RNA-protein correlated and anti-correlated genes (1D Annotation enrichment test, FDR<0.02)

**Supplementary Table S4:** functional enrichments in survival patterns (Fisher Exact test, FDR<0.02)

**Supplementary Table S5:** list of top survival related genes within single cell based signatures

---

**Supplementary Fig. S1:**
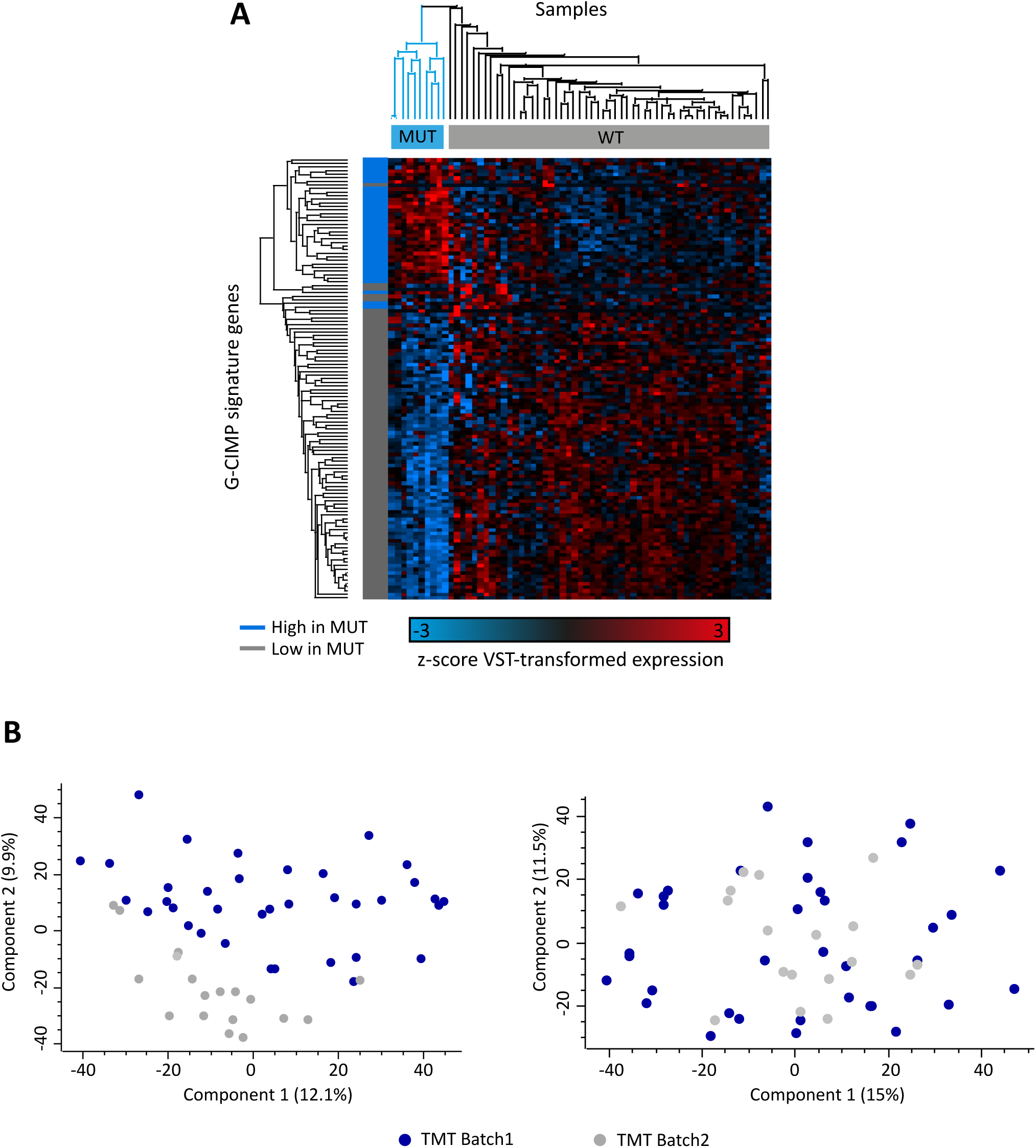
Preprocessing of proteogenomic data. A) Applying Glioma CpG island methylator phenotype (G-CIMP) signature to RNA-seq data to detect IDH-mut samples. Ten samples were identified as mutant, with two of them classified as WT in the clinical information provided (Figure 1C). B) Principal component analysis (PCA) before (left) and after (right) TMT batch effect removal. Different colors represent the two batches of TMT sets.

**Supplementary Fig. S2:**
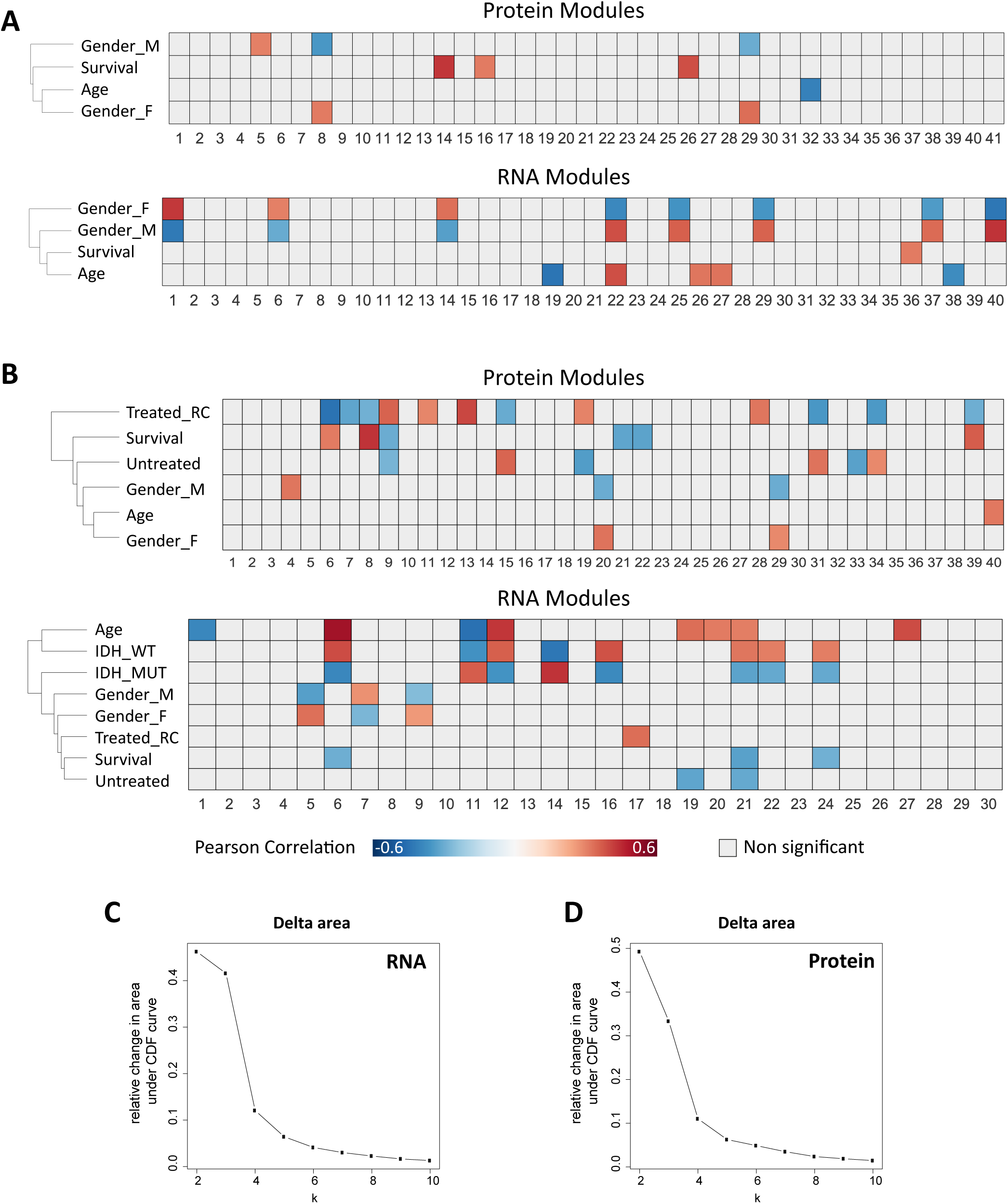
Independent WGCNA and unsupervised clustering of RNA and protein data. A) Comparing protein and RNA modules excluding IDH-mut samples and treated samples. Significantly correlating long-term survival modules are found regaardless of treatment. B) Comparing protein and RNA modules including IDH-mut samples. RNA results show that survival correlating modules are all IDH-correlating as well (Treated_RC=treated with radiation and chemotherapy). Significant Pearsoon correlations (p<0.05) to clinical parameters are indicated in red to blue color, as indicated in the color bar. C+D) Relative change in area under the cumulative distribution function (CDF) curve of consensus scores in RNA (C) and protein (D).

**Supplementary Fig. S3:**
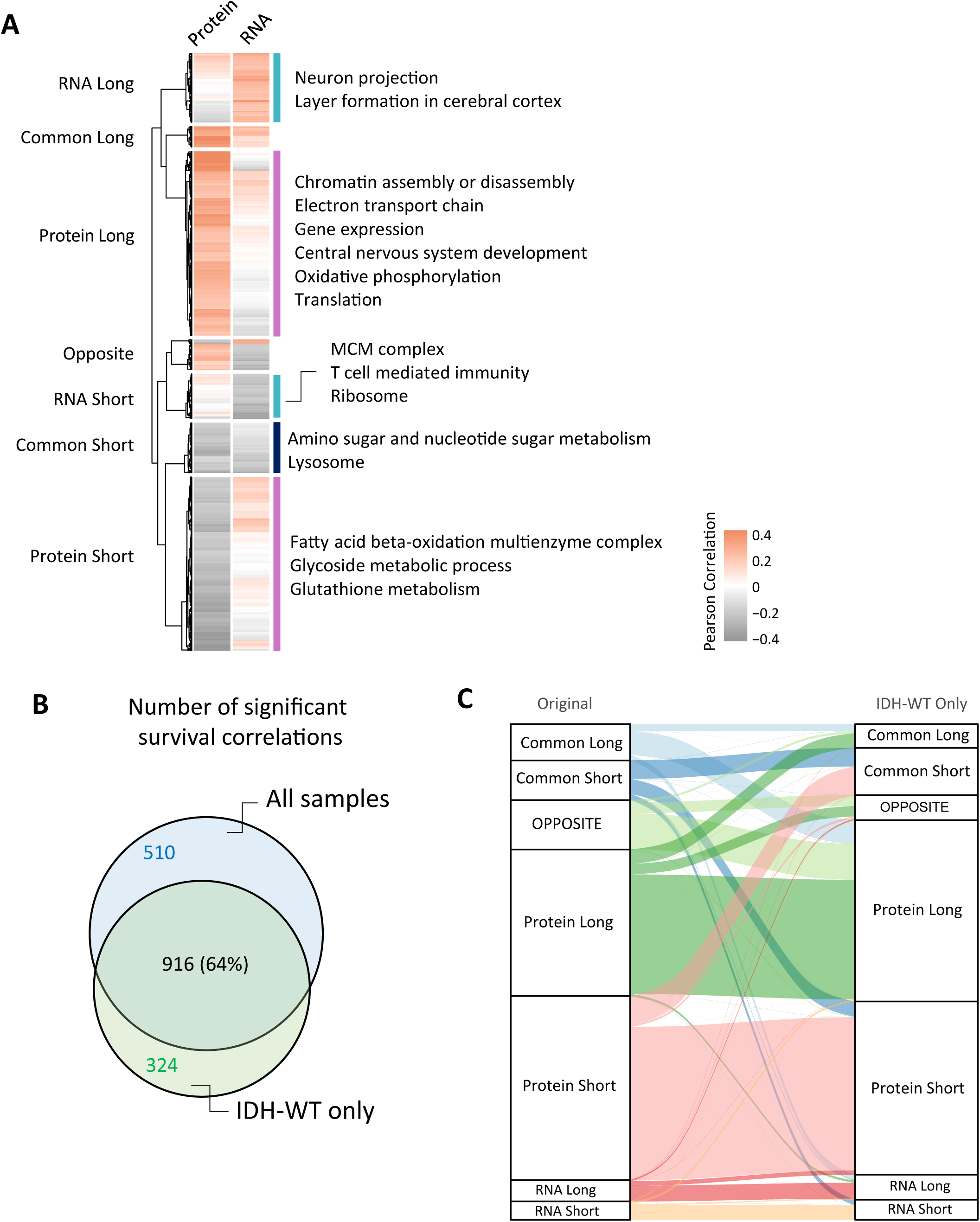
Controlling for IDH mutation - repeating pattern analysis with IDH-WT only. A) Hierarchical clustering of 1240 genes significantly correlating to survival in either RNA or protein (adjusted p-value<0.1) identifies seven clusters. B) Venn diagram showing 64% overlap between the list of 1426 genes in Fig. 4 and the IDH-WT genes. C) Alluvial plot showing the assignment of each of 916 common genes that correlated significantly to survival in both the original analysis and the IDH-WT only. In the layer-specific clusters, most genes belong to the same cluster, while the common and opposite clusters are only partially overlapping.

